# Associations between human milk oligosaccharide concentrations and child growth in a Nicaraguan birth cohort

**DOI:** 10.64898/2026.01.10.26343839

**Authors:** Luis E. Zambrana, Samuel Vilchez, Rebecca J. Rubinstein, Nadja A. Vielot, Fredman González, Lester Gutiérrez, Christian Toval-Ruíz, Stephanie M. Engel, Lars Bode, Filemón Bucardo, Jessie K. Edwards, Sylvia Becker-Dreps

**Affiliations:** Department of Epidemiology, University of North Carolina at Chapel Hill, Chapel Hill, North Carolina, United States of America; Department of Family Medicine, University of North Carolina at Chapel Hill, Chapel Hill, NC, United States; Institute for Global Health and Infectious Diseases, University of North Carolina at Chapel Hill, Chapel Hill, NC, United States; Independent Researcher; Centro de Investigación de Enfermedades Tropicales (CIET). Facultad de Microbiología. Universidad de Costa Rica, San José, Costa Rica; Universidad Tecnológica La Salle, ULSA, León, Nicaragua; Department of Pediatrics, Larsson-Rosenquist-Foundation Mother-Milk-Infant Center of Research Excellence (MOMMI CORE), and the Human Milk Institute (HMI), University of California San Diego (UC San Diego), La Jolla, CA, United States; Department of Microbiology and Immunology, University of North Carolina at Chapel Hill, Chapel Hill, NC, United States

## Abstract

Human milk oligosaccharides (HMOs) are bioactive components in human milk that influence infant health, yet their role in growth trajectories remains unclear. This study examined associations between concentrations of individual HMOs and child growth outcomes: length-for-age (LAZ), weight-for-age (WAZ), and weight-for-length (WLZ) z-scores—at 24 months of age in a population-based cohort from León, Nicaragua. Data were analyzed from 295 mother–infant dyads. HMO concentrations in human milk collected one month postpartum were measured using high-performance liquid chromatography with fluorescence detection. Infant growth was assessed at 10–14 days, 6, and 24 months, with z-scores calculated per WHO standards. Associations between HMOs and growth outcomes were estimated using generalized linear regression models: one unweighted model and two weighted by inverse probability of censoring to address potential bias from differences in breastfeeding duration. Models were adjusted for birth weight, infant sex, maternal secretor status, maternal age, maternal education, and household poverty. Growth associations differed by specific HMOs. 3′-sialyllactose (3′SL; β = 0.11, 95% CI: 0.02, 0.21) and difucosyllactose (DFLac; β = 0.04, 95% CI: 0.00, 0.09) were positively associated with LAZ, suggesting a role in promoting linear growth. In contrast, fucosyllacto-N-hexaose (FLNH; β = −0.07, 95% CI: −0.11, −0.01) was negatively associated with WAZ, and disialyllacto-N-hexaose (DSLNH; β = −0.13, 95% CI: −0.21, −0.05) was negatively associated with WLZ, indicating a more complex relationship with weight gain. These associations remained consistent after IPCW adjustment. Distinct HMOs were differentially associated with child growth patterns. The positive associations of 3′SL and DFLac with LAZ and the negative associations of FLNH and DSLNH with WAZ and WLZ underscore the complexity of HMO–growth relationships and highlight the need for future studies to determine whether targeted HMO modulation or supplementation could optimize early growth outcomes.

## Introduction

HMOs are complex carbohydrates that are abundant in human milk and are thought to play an important role in the development and health of infants (1,2). Recent research has highlighted the potential benefits of HMOs during infancy, particularly in terms of enhancing immune function, shaping the gut microbiota, and potentially influencing growth parameters (3–5).

HMOs have diverse chemical structures, consisting of various monosaccharide units that can form over 200 distinct oligosaccharides(2,6). This structural diversity allows HMOs to serve multiple roles, such as promoting the growth of beneficial gut microbiota (7), preventing pathogen adhesion (8), and modulating immune responses (9,10). For instance, the HMOs, 2’-fucosyllactose (2’-FL) and lacto-N-neotetraose (LNnT), can foster the growth of *Bifidobacterium* species, which are associated with healthy infant gut development and immune system maturation (11).

Recent evidence reveals a range of associations between specific HMOs and child growth metrics, including Length-for-Age z-score (LAZ), Weight-for-Age z-score (WAZ), and Weight-for-Length z-score (WLZ). For example, positive associations have been observed between 2’-FL concentrations and LAZ and WAZ in early childhood, suggesting a beneficial role in promoting linear growth and weight gain (4,12,13).

Conversely, other HMOs, such as LNnT have been negatively associated with these growth metrics; higher concentrations were linked to lower LAZ and WAZ (14–16).

Interestingly, the effects of HMOs on growth may vary among children based on various host factors (17,18), including their geographic location(19,20), as gut microbiome composition differs across geographic regions. These variations may result from local dietary patterns, exposure to region-specific pathogens and microbes, or the overall HMO profile in human milk, which is partly influenced by maternal genetics.

Additionally, fluctuations in HMO concentrations may reflect maternal nutritional status and environmental stressors, all of which play a role in infant growth and health outcomes (21,22).

Maternal secretor status, ascertained by the presence or absence of functional FUT2 gene expression, plays a crucial role in shaping the composition of HMOs in human milk. This, in turn, influences infant gut microbiota development, immune function, and susceptibility to infections, making it a key factor in early-life nutrition and health outcomes (23,24).

Also, differential effects may be associated with residence in high-income countries vs. low- and middle-income countries (LMICs) (21). In LMICs, where chronic malnutrition and infectious diseases are more common, HMOs may offer substantial benefits by promoting immune system development and growth (16,25). In contrast, in high-income countries where overnutrition may be a concern, HMOs could potentially contribute to greater weight gain (12,15,16). This geographic variability underscores the importance of research on the effects of HMOs in diverse populations.

This population-based cohort study in León, Nicaragua, examined the association between specific HMOs and growth metrics at 24 months of age. This timepoint near the end of the “first 1000 days” was selected because it marks a significant milestone that reflects the cumulative impacts of early life nutrition and has profound implications for the child’s future physical growth and cognitive trajectories (26).

We measured the predominant HMOs in human milk samples collected one month postpartum and evaluated their relationship with infant growth at 24 months of age. Identifying specific HMOs that influence child growth could significantly impact breastfeeding practices and recommendations, guide formula development, and inform nutritional interventions to optimize child health outcomes. Our findings contribute to the understanding of how early nutrition influences growth and development, potentially shaping future interventional studies on nutritional supplementation and infant formula composition.

## Methods

### Study design, eligibility criteria, and ethical approvals

This study utilizes data and samples from the Sapovirus-Associated Gastro-Enteritis (SAGE) birth cohort (“Cohort”), consisting of 444 mother-infant pairs in León, Nicaragua(27). The concentrations of 19 HMOs were measured in human milk samples collected one month postpartum, and anthropometrics were measured in children at enrollment (10-14 days of life), 6 and 24 months of age.

The recruitment period spanned from June 12, 2017, to July 31, 2018. During this time, mothers of all live-born singletons newborn residing in 14 health sectors within the Perla Maria Norori Health District were invited to participate in the Cohort. To be eligible for inclusion in the cohort, mothers needed to have a live singleton infant without severe obstetric complications, reside in the municipality of León, Nicaragua, and be over 15 years of age. (For mothers aged 15 to 17 years of age, informed consent was provided by both the mother and the minor mother’s parent.) (27) Eligibility criteria for newborns included no known immune disorders, history of blood transfusion, or chronic health conditions, gestational age of at least 34 weeks, and birth weight of 2,000 grams or more. Families were included if they did not plan to move within the next year and did not have another child enrolled in the Cohort, ensuring a stable residence and a diverse participant pool. We excluded 25 mother-infant dyads in which the mother was not lactating or did not provide a sufficient human milk sample for HMO concentration analysis. Additionally, 14 children were excluded due to withdrawal from the study before baseline anthropometric measurements, and 86 children were excluded because they discontinued participation before the 24-month anthropometric assessment.

The protocol used in this study was reviewed and approved by the Ethical Committee for Biomedical Research of the Universidad Nacional Autonoma de Nicaragua UNAN-León (Acta No.2 2017) and the University of North Carolina at Chapel Hill Institutional Review Board (Study #: 16-2079, approved 9/01/2016). Written informed consent was collected from mothers and the parents of all children and minor mothers in this study.

### Child anthropometric measurements

Field nurses were trained during the study’s preparatory phase to obtain anthropometric measures on the infants by pediatricians, following standardized protocols. Monthly standardization sessions of the scales were conducted and once a year, an anthropometry expert visited sites to participate in these sessions and refresh the training. In the current analysis, we used birthweight as reported by the delivering hospital and calculated anthropometric z-scores based on the WHO anthropometric standards at the initial visit (baseline anthropometric measures) based on weight and length measurements at 10-14 days after birth and at 24 months. Weight measurements were taken using a Salter weighing scale, which allowed precise readings recorded to the nearest 100 grams. Recumbent length was measured at both timepoints using a standard infantometer(28).

### Covariates

During the initial household visit at 10-14 days after birth, trained fieldworkers collected information on each mother-child dyad, including child sex, birthweight (from the delivery record), gestational age, mode of delivery (vaginal or Cesarean section), maternal age, breastfeeding duration in weeks, and mother’s educational attainment.

Family and household characteristics were used to generate a validated poverty index as described previously(29). Subsequently, fieldworkers visited children in their household every week and assessed for breastfeeding duration. We defined the duration of breastfeeding as the total number of weeks a mother reported breastfeeding her child, including either exclusive or non-exclusive breastfeeding (refers to when an infant receives breast milk along with other liquids or foods) (27).

### Human milk data and oligosaccharides analysis

The Bode Laboratory at the University of California San Diego extracted, identified, and measured the concentrations of 19 unique HMOs in human milk samples collected one-month post-partum using High-Performance Liquid Chromatography with Fluorescence Detection (HPLC-FL) (30). We selected a one-month time point as milk composition in early life is important for gut microbiome establishment(31,32) and it is past the immediate post-partum period where rapid changes in HMOs concentration occur(30,33). Prior studies show that after one month postpartum, the composition of HMO remains relatively constant for some HMOs such as 2’FL and LSTb, though others continue to vary throughout the lactation period (33,34).

The concentrations of the following HMOs (in μg/mL) were determined: 3’-Sialyllactose (3’SL), 2’-Fucosyllactose (2’FL), 3-Fucosyllactose (3FL), Difucosyllactose (DFLac), 6’-Sialyllactose (6’SL), Lacto-N-tetraose (LNT), Lacto-N-neotetraose (LNnT), Lacto-N-fucopentaose I (LNFP-I), Lacto-N-fucopentaose II (LNFP-II), Lacto-N-fucopentaose III (LNFP-III), Sialyl-lacto-N-tetraose b (LSTb), Sialyl-lacto-N-tetraose c (LSTc), Difucosyllacto-N-tetraose (DFLNT), Lacto-N-hexaose (LNH), Disialyllacto-N-tetraose (DSLNT), Fucosyllacto-N-hexaose (FLNH), Difucosyllacto-N-hexaose (DFLNH), Fucosyl-disialyllacto-N-hexaose (FDSLNH), Disialyllacto-N-hexaose (DSLNH).

### Assessment of secretor status

Secretor status was determined in mothers and infants by using a saliva-based assay. In brief, ELISA plates were coated with children’s saliva diluted in PBS, and the presence of the α1,2 fucose residuals was detected by the *Ulex europaeus* lectin peroxidase (UEA-I, Sigma-Aldrich) (35).

### Statistical analysis

Statistical analyses were performed using SAS software, version 9.4 (SAS Institute Inc., Cary, NC)(36). Descriptive statistics, including means, standard deviations, range, and frequencies, were calculated to summarize the demographic and clinical characteristics of the study population, including weight and length measurements, and HMO concentrations. A series of multivariable linear regression analyses using generalized linear models (GLMs) evaluated the associations between each HMO and child growth metrics (LAZ, WAZ, and WLZ at 24 months). Specifically, we examined three primary dependent variables: LAZ-scores at 24 months, WAZ-scores at 24 months, and WLZ-scores at 24 months. Each outcome variable was modeled separately for each HMO using a generalized linear model. The primary parameter of interest was the estimated difference in each growth outcome (e.g., LAZ) associated with an increase in the concentration of each HMO, as obtained from the regression model. To facilitate interpretability and numerical stability, all HMO concentrations (µg/mL) were scaled by multiplying by 0.01 prior to analysis. As a result, the reported regression coefficients represent the expected change in the outcome for a 100 µg/mL increase in HMO concentration, holding all other covariates constant.

All models were adjusted for key covariates, including infant sex (classified with male as the reference), maternal age group (categorized as 15-17, 18-25, 26-35 [reference], and 36+ years), birth weight (grams), maternal secretor status (classified as 0 for non-secretors and 1 [reference] for secretors), maternal education level (categorized as 1 for less than 8 years of education, 2 [reference] for 8 to 12 years of education, and 3 for more than 12 years of education), household poverty status (categorized as 0 [reference] for non-poor and 1 for poor).

In the models for LAZ-score and WLZ-score at 24 months, the baseline value of the outcome (measured at 10-14 days after birth) was included as a covariate. All statistical tests were two-sided. The Wald test was used to evaluate the statistical significance of associations, with significance set at p < 0.05 and evaluation of 95% CI.

In the multivariable regression analysis, we conducted several key diagnostic checks to ensure the validity and reliability of our results. Firstly, we assessed the linearity of the relationship between the independent variables and the dependent variable through visual inspection of scatter plots and residual plots. The normality of the residuals was evaluated using histograms, Q-Q plots, and the Shapiro-Wilk test.

Homoscedasticity was checked by examining the residuals versus fitted values plot. We assessed multicollinearity among the predictor variables using Variance Inflation Factors (VIFs), ensuring no VIF exceeded the threshold of 10, indicating problematic multicollinearity.

Maternal age was explored using multiple functional forms, including linear, quadratic, binary, categorical, and cubic spline representations. After evaluating the model fit, the categorical form (15-17, 18-25, 26-35, 36+ years) provided the best fit and was selected for the final model. We also tested interaction terms between individual HMOs and breastfeeding duration, but these terms did not significantly improve the model’s capacity to capture the relationships between the exposures and the child growth outcomes. Consequently, interaction terms were not included in the final model.

The primary analysis was conducted using complete case analysis (CCA), where observations with missing values in the household poverty status variable (n = 24; 7.5%) were excluded.

Because many HMOs are structurally and biologically correlated, applying strict multiple-comparison corrections such as Bonferroni would have substantially reduced statistical power and increased the likelihood of Type II errors. Therefore, given the exploratory and hypothesis-generating nature of this study, results were interpreted based on effect sizes, confidence intervals, and overall consistency rather than on strict significance thresholds.

### Analysis accounting for duration of breastfeeding

Given that breastfeeding duration is a key predictor of the outcome, we conducted a second set of analyses that accounted for differences in breastfeeding duration. We censored participants who did not breastfeed for 6 months (analysis 1) and who did not breastfeed for the full 24-month period (analysis 2). To address potential selection bias introduced by this exclusion, we applied IPCW, which helps correct for systematic differences between those who continued breastfeeding and those who did not. Logistic regression models were used to estimate the probability of breastfeeding continuation at two key points: 6 months and 24 months. The weights were calculated as the inverse of these probabilities, ensuring that individuals with lower probabilities of continued breastfeeding were appropriately weighted. Additionally, we stabilized the weights to improve efficiency and reduce variability in the estimates. For example, the 6-month IPCW model examines the associations in a hypothetical population where all children breastfed for 6 months, while the 24-month IPCW model examined the associations in a hypothetical population where all children breastfed for 24-months. This approach ensures that our findings reflect the full population rather than being biased by the exclusion of infants who stopped breastfeeding.

### Sensitivity Analysis

To assess the robustness of our findings, we conducted sensitivity analyses incorporating HMO group classifications—Sialylated, Fucosylated, and Non-Fucosylated—as covariates in our models. This approach allowed us to evaluate whether the observed associations between individual HMOs and the outcome variable remained consistent when accounting for the broader structural categories of HMOs.

Before conducting the sensitivity analyses, we assessed potential multicollinearity among individual HMO concentrations using a correlation matrix. Most pairwise correlations were low to moderate; however, higher correlations were observed among HMOs within the same structural groups and some among different HMO groups. This pattern reflects the biological relationships among HMOs and suggests that some degree of multicollinearity may be present within HMO groups.

### Residual Adjustment (residual centering) for Multicollinearity

To ensure that the observed associations reflect the independent contribution of each individual HMO, we applied residual centering to account for shared variance among the HMO variables. Specifically, each HMO group variable was regressed on the index HMO under investigation, effectively partitioning the variance shared between the index HMO and the broader HMO group. The resulting residuals, which represent the variance in each group variable that is not explained by the index HMO, were then included in the regression models (37).

While this approach does not completely eliminate multicollinearity, it effectively reduces the direct influence of correlations within HMO groups, allowing for a clearer interpretation of each HMO’s unique effect.

## Results

### Study Population and Demographic Characteristics

The SAGE study enrolled 444 mother-child dyads, as shown in **Fig 1**. After excluding 25 mothers who were not lactating or did not provide milk samples, 419 dyads remained for HMO analysis. An additional 14 children were lost to follow-up before baseline anthropometric measurements, reducing the sample to 405. By the 24-month follow-up, 86 more children had dropped out, leaving 319 dyads. Of these, 24 were excluded due to missing Household Poverty Status data, resulting in a final analytical sample of 295.

**Fig 1.**
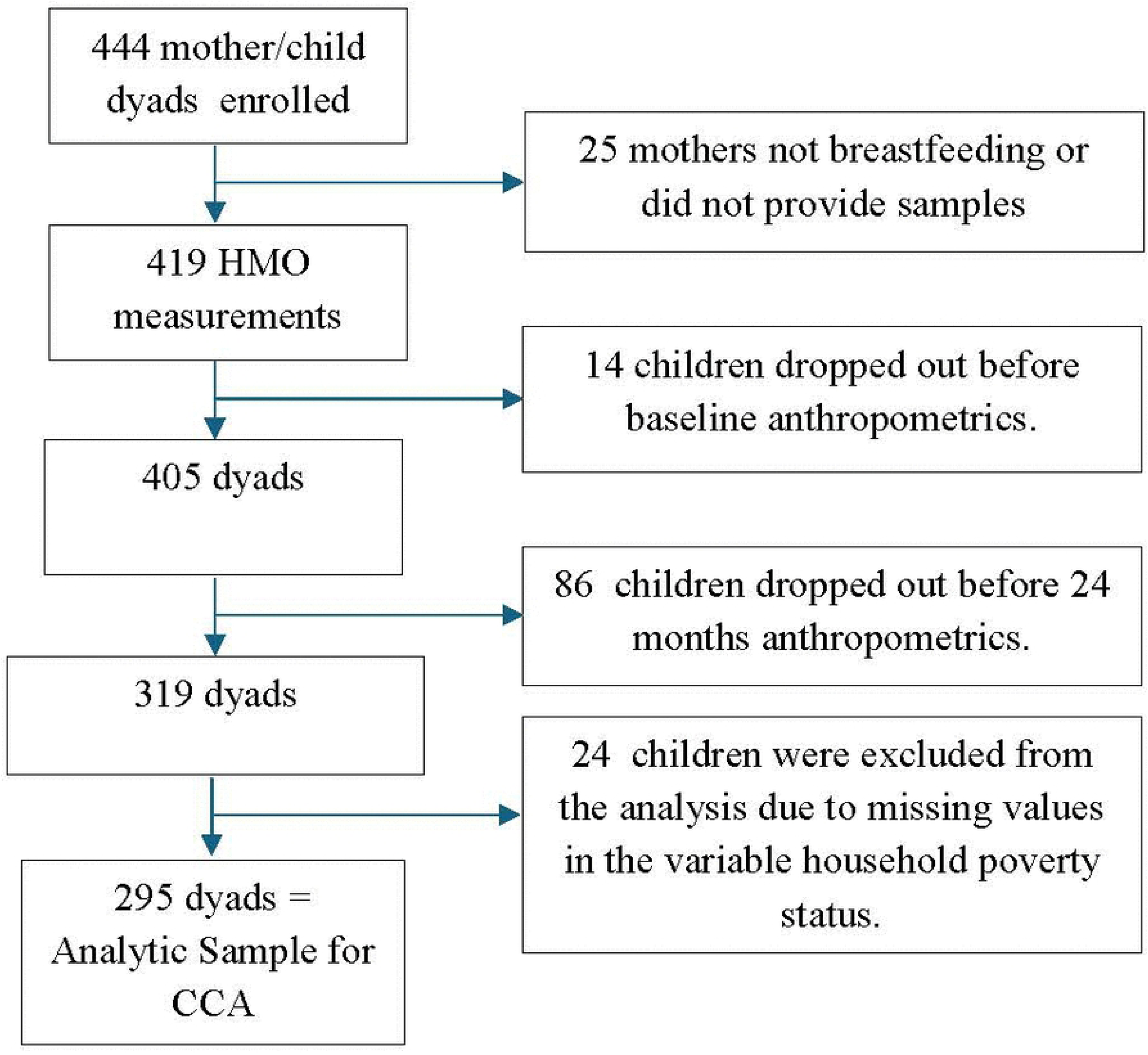
Flow diagram of mother–child dyads included in the study. A total of 444 mother–child dyads were enrolled. Among them, 25 mothers were not lactating or did not provide milk samples, resulting in 419 dyads with available human milk oligosaccharide (HMO) measurements. Fourteen children dropped out before baseline anthropometric assessment, reducing the sample to 405 dyads. An additional 86 children dropped out before the 24-month visit, leaving 319 dyads. Of these, 24 dyads were excluded due to missing household poverty data, yielding a final analytic sample of 295 dyads for complete case analysis.

The demographic characteristics of the study population are detailed in **Table 1**. The mean maternal age was 24.2 years (SD ± 5.0), with mothers having more than 12 years of education (50.2%) and working in informal jobs (85.8%). The children in the study were nearly evenly split by sex, with 50.8% male and 49.2% female. The vast majority of mothers (89.8%) were secretors, a key determinant in the variation of HMO concentrations in human milk.

**Table 1.**
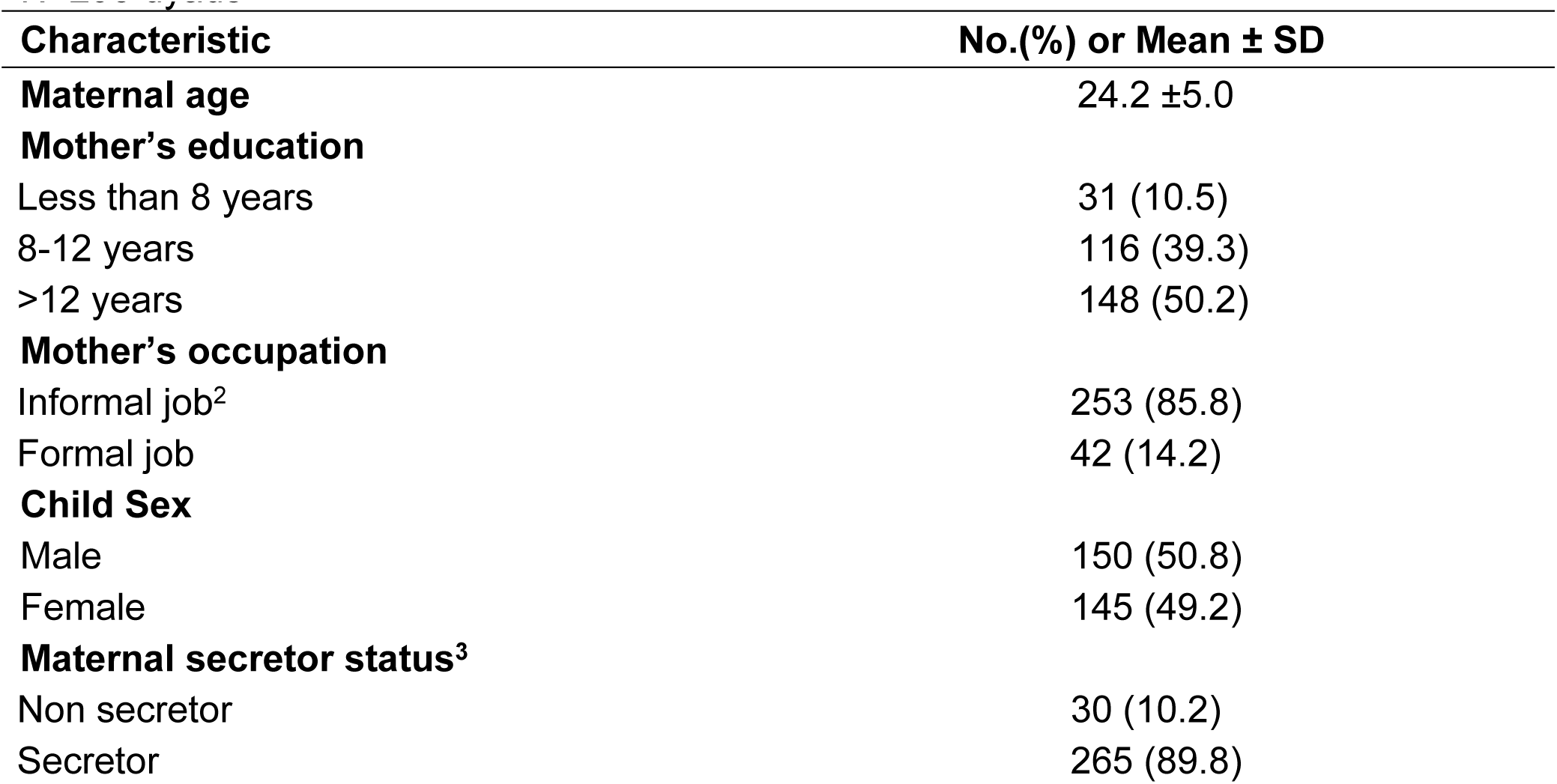
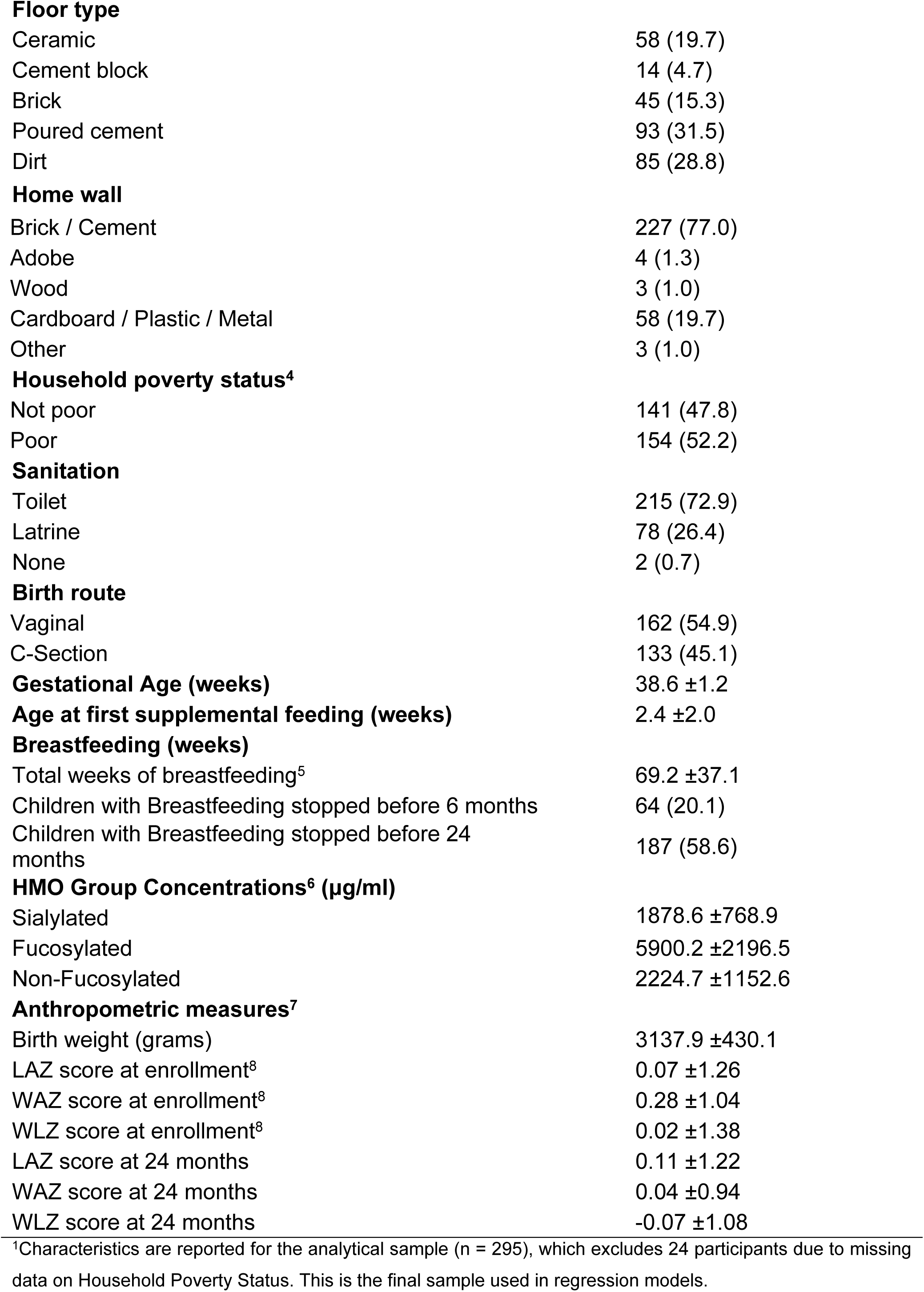
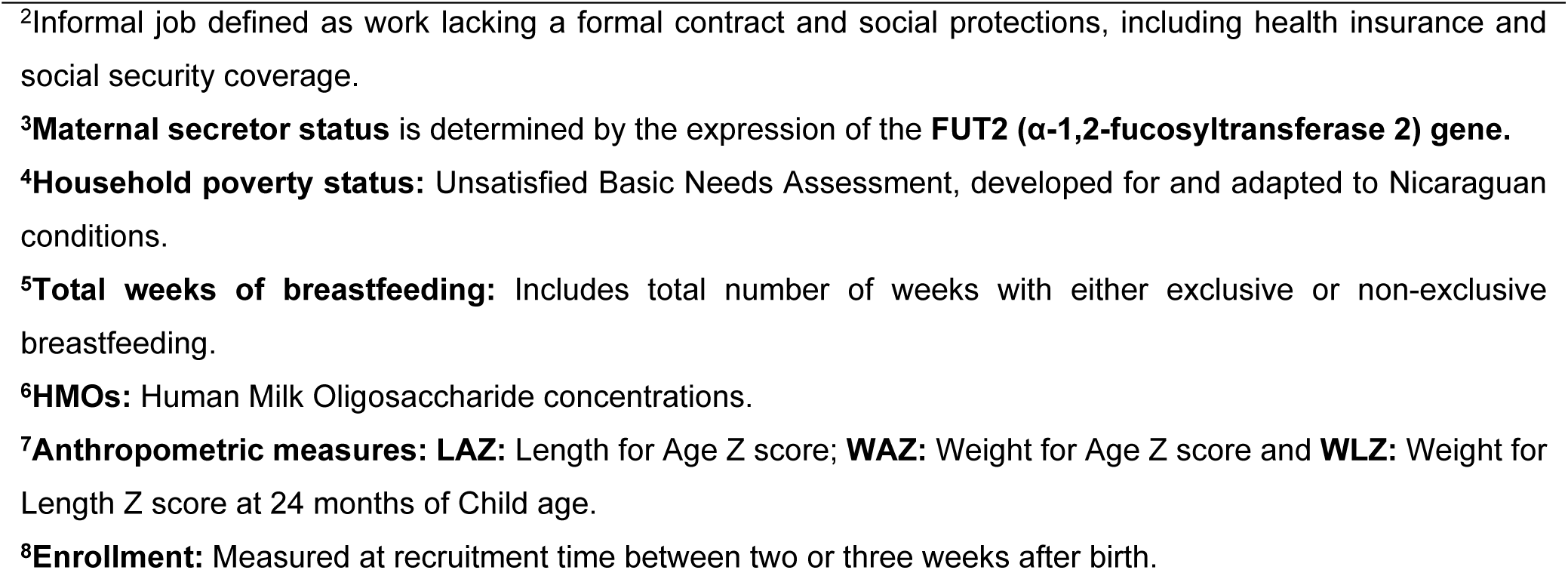
Demographic characteristics of mother-infant dyads and key study variables. N=295 dyads^1^.

| Characteristic | No.(%) or Mean ± SD |
| --- | --- |
| **Maternal age** | 24.2 ± 5.0 |
| **Mother's education** |  |
| Less than 8 years | 31 (10.5) |
| 8-12 years | 116 (39.3) |
| >12 years | 148 (50.2) |
| **Mother's occupation** |  |
| Informal job<sup>2</sup> | 253 (85.8) |
| Formal job | 42 (14.2) |
| **Child Sex** |  |
| Male | 150 (50.8) |
| Female | 145 (49.2) |
| **Maternal secretor status<sup>3</sup>** |  |
| Non secretor | 30 (10.2) |
| Secretor | 265 (89.8) |
| **Floor type** |  |
| Ceramic | 58 (19.7) |
| Cement block | 14 (4.7) |
| Brick | 45 (15.3) |
| Poured cement | 93 (31.5) |
| Dirt | 85 (28.8) |
| **Home wall** |  |
| Brick / Cement | 227 (77.0) |
| Adobe | 4 (1.3) |
| Wood | 3 (1.0) |
| Cardboard / Plastic / Metal | 58 (19.7) |
| Other | 3 (1.0) |
| **Household poverty status<sup>4</sup>** |  |
| Not poor | 141 (47.8) |
| Poor | 154 (52.2) |
| **Sanitation** |  |
| Toilet | 215 (72.9) |
| Latrine | 78 (26.4) |
| None | 2 (0.7) |
| **Birth route** |  |
| Vaginal | 162 (54.9) |
| C-Section | 133 (45.1) |
| **Gestational Age (weeks)** | 38.6 ±1.2 |
| **Age at first supplemental feeding (weeks)** | 2.4 ±2.0 |
| **Breastfeeding (weeks)** |  |
| Total weeks of breastfeeding<sup>5</sup> | 69.2 ±37.1 |
| Children with Breastfeeding stopped before 6 months | 64 (20.1) |
| Children with Breastfeeding stopped before 24 months | 187 (58.6) |
| **HMO Group Concentrations<sup>6</sup> (µg/ml)** |  |
| Sialylated | 1878.6 ±768.9 |
| Fucosylated | 5900.2 ±2196.5 |
| Non-Fucosylated | 2224.7 ±1152.6 |
| **Anthropometric measures<sup>7</sup>** |  |
| Birth weight (grams) | 3137.9 ±430.1 |
| LAZ score at enrollment<sup>8</sup> | 0.07 ±1.26 |
| WAZ score at enrollment<sup>8</sup> | 0.28 ±1.04 |
| WLZ score at enrollment<sup>8</sup> | 0.02 ±1.38 |
| LAZ score at 24 months | 0.11 ±1.22 |
| WAZ score at 24 months | 0.04 ±0.94 |
| WLZ score at 24 months | -0.07 ±1.08 |

<sup>1</sup>Characteristics are reported for the analytical sample (n = 295), which excludes 24 participants due to missing data on Household Poverty Status. This is the final sample used in regression models.
<sup>2</sup>Informal job defined as work lacking a formal contract and social protections, including health insurance and social security coverage.
<sup>3</sup>**Maternal secretor status** is determined by the expression of the **FUT2 ( $\alpha$ -1,2-fucosyltransferase 2) gene**.
<sup>4</sup>**Household poverty status**: Unsatisfied Basic Needs Assessment, developed for and adapted to Nicaraguan conditions.
<sup>5</sup>**Total weeks of breastfeeding**: Includes total number of weeks with either exclusive or non-exclusive breastfeeding.
<sup>6</sup>**HMOs**: Human Milk Oligosaccharide concentrations.
<sup>7</sup>**Anthropometric measures**: **LAZ**: Length for Age Z score; **WAZ**: Weight for Age Z score and **WLZ**: Weight for Length Z score at 24 months of Child age.
<sup>8</sup>**Enrollment**: Measured at recruitment time between two or three weeks after birth.

### HMO Concentrations

The median concentrations of key HMOs stratified by maternal secretor status are presented in **Table 2**. Out of the 19 HMOs measured, only the concentrations of LNT, 3FL, DSLNH, LNnT, LNH, and LNFP-III did not differ significantly by secretor status. The predominant HMOs in the maternal secretor were 2’FL and LNFP-I, with median concentrations of 2,916.6 µg/mL [Q1: 2001.1, Q3: 4149.9] and 1029.5 µg/mL [Q1: 567.24; Q3: 1604.4], respectively. In contrast, the most abundant HMOs in non-secretor mothers were LNT (1232.7 µg/mL [Q1: 603.2, Q3: 1828.9]) and LNFP-II (1159.7 µg/mL [Q1:732.4, Q3: 1468.6]). The least abundant HMO in secretor mothers is LNFP-III (46.4 µg/mL [Q1:29.2, Q3: 76.5]), and in non-secretor mothers is 2’FL (7.8 µg/mL [Q1:4.2, Q3: 13.8]).

**Table 2.** Descriptive statistics of the HMOs stratified by secretor status and overall given as median [Q1; Q3]^1^.

| HMO (µg/mL) | Total (n=319) | Secretor mother (n=289) | Non-secretor mother (n=30) | p-value<sup>2</sup> |
| --- | --- | --- | --- | --- |
| 2'FL | 2682.3 [1691.2, 4034.4] | 2916.6 [2001.1, 4149.9] | 7.8 [4.2, 13.8] | 0.001 |
| LNFP_I | 914.9 [471.1, 1536.4] | 1029.5 [567.2, 1604.3] | 173.5 [131.6, 229.3] | 0.001 |
| DFLNT | 884.9 [416.3, 1476.1] | 968.6 [519.6, 1572.1] | 343.9 [144.3, 457.4] | 0.001 |
| LNT | 612.3 [375.1, 955.7] | 596.3 [370.9, 888.8] | 1232.7 [603.2, 1828.9] | 0.16 |
| LNFP_II | 583.7 [379.4, 852.9] | 562.3 [374.3, 815.3] | 1159.7 [732.4, 1468.6] | 0.045 |
| 6'SL | 504.1 [334.4, 701.2] | 497.5 [330.0, 694.6] | 626.8 [379.6, 887.1] | 0.001 |
| 3FL | 404.6 [242.8, 671.4] | 375.7 [234.3, 587.0] | 1119.4 [576.5, 1583.2] | 0.068 |
| FLNH | 284.8 [184.7, 390.0] | 275.6 [177.9, 377.2] | 347.9 [264.7, 661.3] | 0.001 |
| LSTc | 281.1 [188.5, 388.6] | 285.3 [196.8, 403.5] | 207.6 [152.1, 312.7] | 0.001 |
| DSLNH | 224.2 [144.3, 332.9] | 224.2 [143.4, 331.7] | 217.8 [162.5, 354.0] | 0.62 |
| DFLac | 232.6 [133.1, 395.7] | 250.3 [163.5, 439.2] | 22.9 [14.0, 28.2] | 0.016 |
| FDSLNH | 209.0 [134.9, 391.0] | 193.8 [132.1, 316.7] | 500.5 [280.9, 758.8] | 0.002 |
| DSLNT | 171.9 [113.8, 262.7] | 174.1 [113.9, 259.4] | 167.1 [112.3, 287.0] | 0.001 |
| LNnT | 145.8 [97.6, 218.2] | 151.7 [99.9, 219.1] | 126.8 [72.1, 189.1] | 0.81 |
| LNH | 147.6 [98.3, 220.8] | 148.4 [98.9, 220.8] | 136.0 [97.9, 218.6] | 0.98 |
| 3'SL | 113.1 [82.9, 169.2] | 113.8 [84.6, 175.4] | 105.7 [76.1, 127.8] | 0.002 |
| LSTb | 88.5 [60.0, 133.3] | 88.2 [57.8, 129.3] | 110.2 [71.2, 173.8] | 0.001 |
| DFLNH | 85.2 [47.3, 155.1] | 82.2 [46.4, 138.3] | 205.9 [122.5, 293.3] | 0.001 |
| LNFP_III | 46.5 [28.8, 76.2] | 46.4 [29.2, 76.5] | 48.8 [20.2, 71.2] | 0.72 |

<sup>1</sup>Descriptive statistics are based on the full study sample (n = 319) to provide an unbiased summary of HMO concentrations.
<sup>2</sup>Group differences were tested using Kruskal-Wallis-tests.
Abbreviations: 2'FL, 2-fucosyllactose; 3FL, 3-fucosyllactose; DFLac, difucosyllactose; 3'SL, 3-sialyllactose; 6'SL, 6-sialyllactose; LNT, lacto-N-tetrose; LNnT, lacto-N-neotetraose; LNFP I, lacto-N-fucopentaose I; LNFP II, lacto-N-fucopentaose II; LNFP III, lacto-N-fucopentaose III; LSTb, sialyl-lacto-N-tetraose b; LSTc, sialyl-lacto-N-tetraose c; DFLNT, difucosyllacto-N-tetraose; LNH, lacto-N-hexaose; DSLNT, disialyllacto-N-tetraose; FLNH, fucosyllacto-N-hexaose; DFLNH, difucosyllacto-N-hexaose; FDSLNH, fucodisialyllacto-N-hexaose; DSLNH, disialyllacto-N-hexaose.

### HMO Concentrations and Infant Growth Outcomes

This section presents the analysis using CCA, which included 295 observations with complete data. To facilitate the interpretation of regression coefficients, concentrations of HMOs were multiplied by 0.01 before modeling (beta coefficients from the scaled model represent a change in the outcome per 100 units of the predictor). The associations between HMOs and infant growth outcomes at 24 months were evaluated using forest plots, which depict the effect sizes and 95% confidence intervals (CIs) for each HMO in relation to three growth outcomes: LAZ, WAZ, and WLZ. The HMOs were organized into three categories—sialylated, fucosylated, and non-fucosylated—each distinguished by different colors in the plots. All models were adjusted for key covariates, including infant sex, maternal age group, birth weight, maternal secretor status, maternal education, and household poverty status.

Overall, the results show that specific HMOs, particularly those in the sialylated group, are more likely to be positively associated with higher LAZ. However, the significance and direction of these associations varied between different HMOs and across growth measures.

**Fig 2**, Panel A illustrates the adjusted associations between individual HMOs and LAZ at 24 months, accounting for key covariates. Among the analyzed HMOs, several sialylated and fucosylated HMOs showed positive associations with LAZ.

**Fig 2.**
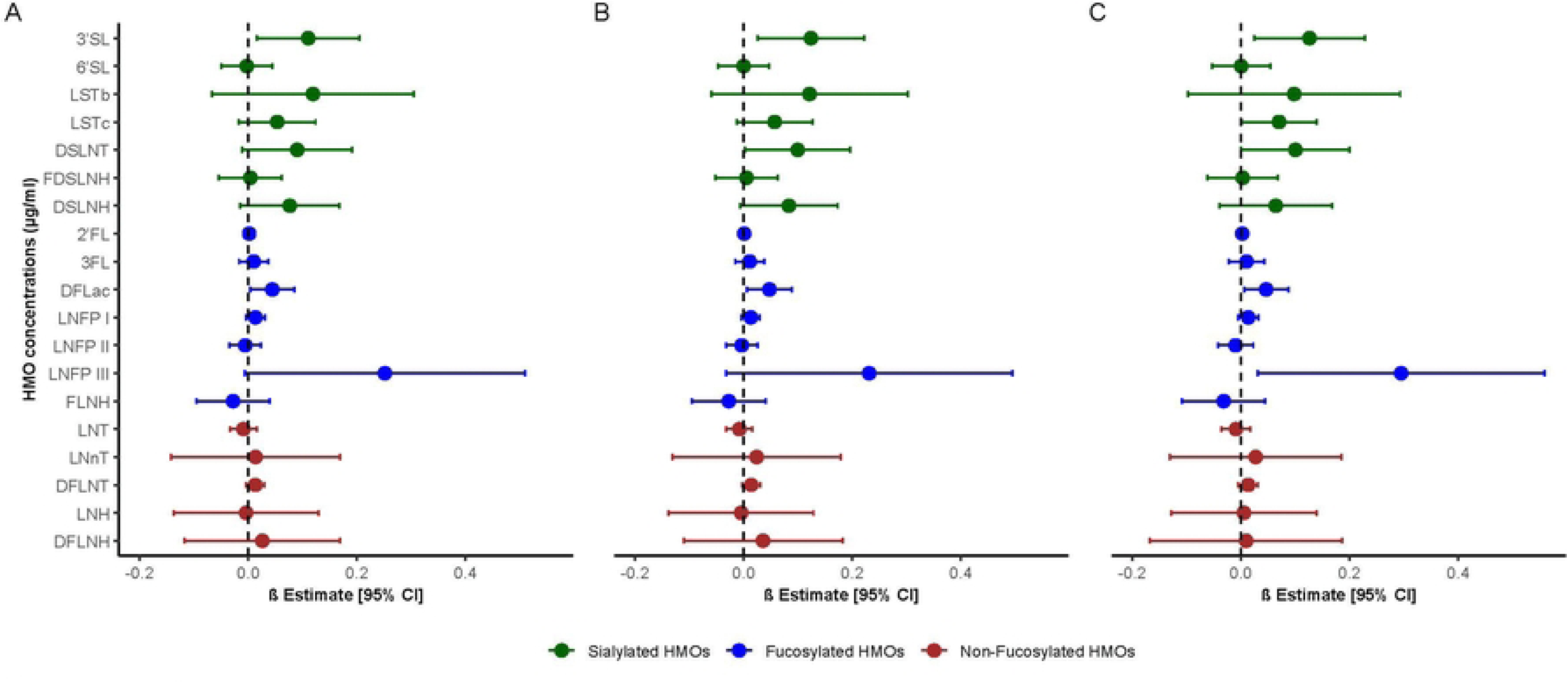
Associations between human milk oligosaccharide concentrations at one month postpartum and length-for-age z-scores at 24 months. Data are presented as β coefficients and 95% confidence intervals from multivariable linear regression models. (A) Unweighted model. (B) Model weighted using inverse probability of censoring weights (IPCW) assuming all participants were breastfed at 6 months. (C) IPCW-adjusted model assuming breastfeeding for 24 months. Sialylated HMOs (green), fucosylated HMOs (blue), and non-fucosylated HMOs (red) are shown. Models were adjusted for baseline LAZ, infant sex, maternal age, birth weight, maternal secretor status (defined by the presence or absence of 2′-fucosyllactose), maternal education, and household poverty. HMO concentrations were scaled by multiplying by 0.01; β coefficients represent the change in the outcome per 100-unit increase in the predictor.

3’SL was positively associated with LAZ such that for every 100 μg/mL increase in 3’SL, the LAZ score increased by 0.11 (95% CI: 0.02, 0.21) on average. DFLac also showed a positive association, with 0.04 (95% CI: 0.00, 0.09) per 100 μg/mL increase.

This suggests that a 100 μg/mL increase in DFLac was associated with a 0.04 z-score unit increase in LAZ. Other sialylated HMOs, including LSTb, LSTc, DSLNT, and DSLNH, also exhibited positive associations with LAZ, although with confidence intervals crossing the null.

Fig 2, panels B and C present weighted estimates assuming all participants breastfed for 6 and 24 months, respectively. Patterns across HMO groups were largely consistent, with minor variations in estimates and CIs. 3’SL and DFLac remained positively associated with growth, and after weighting for 24 months of breastfeeding, the 95% CIs for LSTc and DSLNT no longer included the null.

**Fig 3** illustrates the association between HMOs at one month postpartum and WAZ at 24 months, comparing the difference in WAZ for each 0.01-unit increase in each HMO and 95% CIs across three models. Panel A presents results from the multivariate model without adjustment for breastfeeding duration. The findings reveal mixed effects among the three HMO groups: while 3’SL, LSTb, and DFLac exhibit weakly positive associations, their CIs include the null value. In contrast, certain HMOs—including the sialylated HMOs, 6’SL and DSLNH, the fucosylated HMO, FLNH, and the non-fucosylated HMO, LNH, demonstrate negative associations with growth.

**Fig 3.**
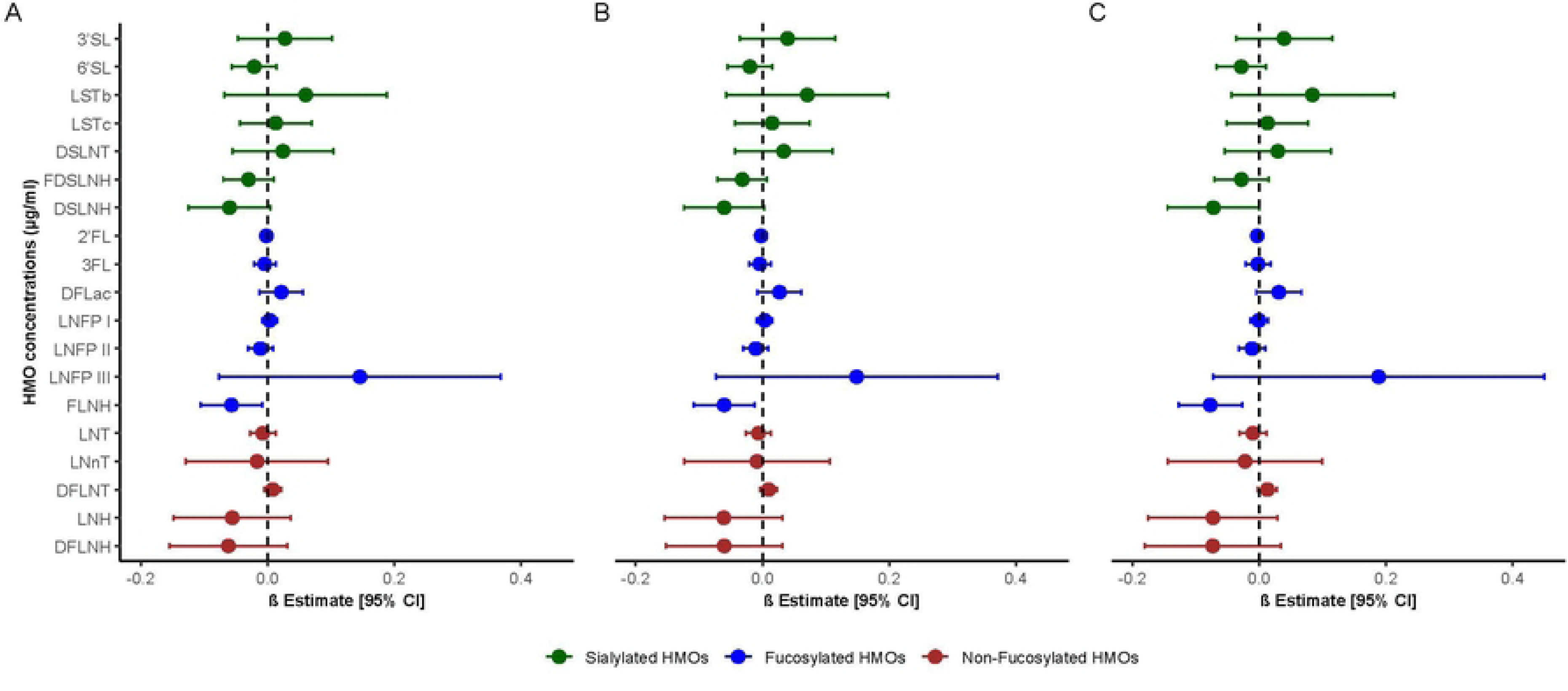
Associations between human milk oligosaccharide concentrations at one month postpartum and weight-for-age z-scores at 24 months. Data are presented as β coefficients and 95% confidence intervals from multivariable linear regression models. (A) Unweighted model. (B) Model weighted using inverse probability of censoring weights (IPCW) assuming all participants were breastfed at 6 months. (C) IPCW-adjusted model assuming breastfeeding for 24 months. Sialylated HMOs (green), fucosylated HMOs (blue), and non-fucosylated HMOs (red) are shown. Models were adjusted for infant sex, maternal age, birth weight, maternal secretor status (presence or absence of 2′-fucosyllactose), maternal education, and household poverty. HMO concentrations were scaled by multiplying by 0.01; β coefficients represent the change in the outcome per 100-unit increase in the predictor.

For example, FLNH exhibits a negative association with WAZ (difference per 0.01-unit change = -0.07, 95% CI: -0.11, -0.01). This relationship indicates that elevated early FLNH levels may correspond to weights modestly below the median for age at two years.

As in Fig 2, Fig 3, Panels B and C present results had all participants continued breastfeeding for 6 and 24 months, respectively. The estimates and their precision remain largely consistent with those in Panel A. Of particular interest, FLNH maintains a negative association with WAZ across all three models, even after applying the weighted analysis, reinforcing a potential inverse relationship between early-life FLNH exposure and later growth outcomes.

**Fig 4** depicts the associations between HMOs at one month postpartum and WLZ at 24 months old, comparing estimates and 95% CIs across the same three scenarios as for LAZ and WAZ. Panel A shows negative associations between the sialylated HMO DSLNH and WLZ at 24 months with estimates -0.13 and 95%CI (-0.21, -0.05). Some other HMOs, such as the fucosylated HMO FLNH, and the non-fucosylated LNH tended to have a negative association. None of the HMOs show a positive association with WLZ at 24 months.

**Fig 4.**
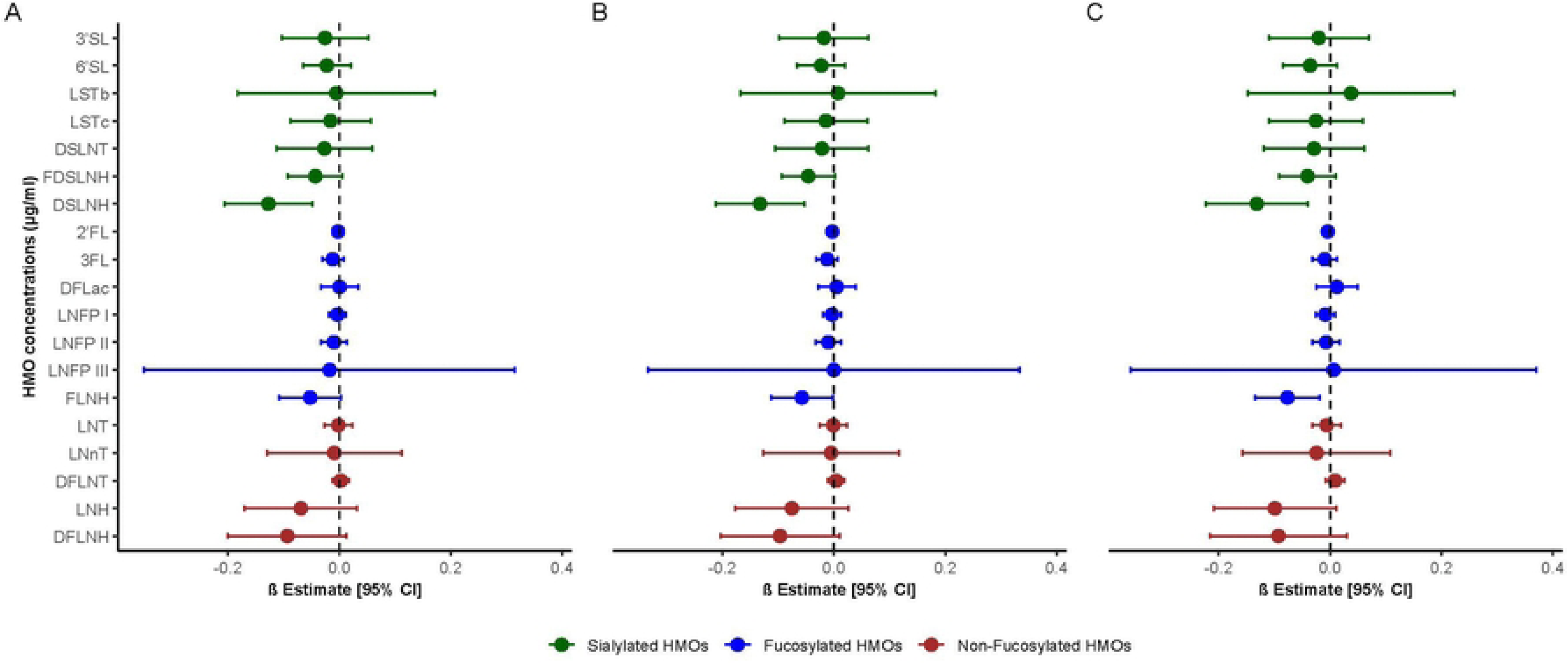
Associations between human milk oligosaccharide concentrations at one month postpartum and weight-for-length z-scores at 24 months. Data are presented as β coefficients and 95% confidence intervals from multivariable linear regression models. (A) Unweighted model. (B) Model weighted using inverse probability of censoring weights (IPCW) assuming all participants were breastfed at 6 months. (C) IPCW-adjusted model assuming breastfeeding for 24 months. Sialylated HMOs (green), fucosylated HMOs (blue), and non-fucosylated HMOs (red) are shown. Models were adjusted for baseline WLZ, infant sex, maternal age, birth weight, maternal secretor status (presence or absence of 2′-fucosyllactose), maternal education, and household poverty. HMO concentrations were scaled by multiplying by 0.01; β coefficients represent the change in the outcome per 100-unit increase in the predictor.

Again, Panels B and C of Figure 4 illustrate models had all participants continued breastfeeding for 6 and 24 months. The patterns observed are generally consistent with those in Panel A. Notably, the sialylated HMO DSLNH and the fucosylated HMO FLNH maintain negative associations at both 6 and 24 months after applying the weighting, reinforcing their inverse association with the outcome.

In summary, the results highlight distinct associations between HMOs at one month postpartum and growth outcomes at 24 months, emphasizing the effect of selected HMOs on early childhood development. Notably, sialylated HMOs such as 3’SL, DFLac, LSTc, and DSLNT demonstrated positive associations with LAZ, with 3’SL and DFLac consistent across both the adjusted multivariate model and the weighted models. Conversely, FLNH and DSLNH exhibited negative associations with WAZ, underscoring potential inverse relationships between these HMOs and WAZ outcomes.

Sensitivity analyses confirmed the robustness of these findings, showing no meaningful differences in estimates or CIs after accounting for possible high HMO group correlation (S Fig).

## Discussion

This study examines the association between specific HMO concentrations at one month postpartum and infant growth metrics at 24 months of age, providing evidence that specific HMOs may predict infant growth trajectories over the first 24 months of life. Sialylated HMOs, particularly 3’SL, DFLac, LSTc, and DSLNT, exhibited a positive relationship with LAZ, supporting a role in promoting linear growth. In contrast, FLNH and DSLNH showed significant negative associations with WAZ, indicating a more complex relationship with weight gain. These associations remained consistent across IPCW-adjusted models and sensitivity analyses, reinforcing the robustness of the findings.

Previous research has yielded mixed findings regarding the relationship between HMOs and infant growth metrics, particularly LAZ. Consistent with our findings, studies conducted in Australia (2022) and Denmark (2019) reported positive associations between 3’SL and LAZ (14,38). Similarly, Larsson et al. (2019) in Denmark identified a positive relationship between DFLac and LAZ (14). In the United States, Alderete et al. (2015) observed a positive association between DSLNT and LAZ, a trend that was also evident in our study (12). In contrast, other studies have reported inverse relationships between HMOs and infant growth outcomes. For instance, Binia et al. (2021) in Germany and Tonon et al. (2019) in Brazil found negative associations between 3’SL and LAZ (23,39). These discrepancies may stem from differences in HMO composition due to maternal characteristics, environmental factors, and methodological variations across studies.

Expanding on these findings, our results further support the role of sialylated and fucosylated HMOs in promoting linear growth during early childhood. The observation that 3’SL, and DFLac were positively associated with LAZ, reinforcing prior research from Australia and Denmark. Additionally, after weighting for breastfeeding duration using IPCW models (Panels B and C), LSTc and DSLNT emerged as significant predictors of LAZ at 24 months, underscoring the potential importance of sialylated HMOs. These results suggest that HMO effects on infant growth may extend beyond individual oligosaccharides, highlighting the synergistic roles of HMO groups in gut microbiota modulation, immune development(5), and nutrient absorption(40). The consistency of associations across different statistical models strengthens the potential causal role of sialylated HMOs in early childhood growth trajectories. However, regional disparities in findings, including the negative associations observed in Germany and Brazil, emphasize the need for mechanistic studies to investigate genetic, dietary, and environmental factors that may influence these relationships.

LNFP-III, a fucosylated HMO, showed a positive association; however, the 95% CI was extremely imprecise. A wide CI suggests high variability or potential imprecision in the estimate, making the observed association unreliable. This may be because it was the least abundant HMO in secretor mothers. Therefore, while the direction of association appears positive, this result should be interpreted with caution until further evidence confirms its reliability.

Regarding WAZ, we identified negative associations with both DSLNH and FLNH. While the inverse relationship between FLNH and WAZ has been previously documented (41), findings related to DSLNH remain inconsistent. Some studies have reported positive associations between DSLNH and WAZ, as well as with other sialylated HMOs such as 3’SL (12). However, our analysis diverged, revealing a negative association between DSLNH and WAZ, a relationship that, to our knowledge, has not been previously reported. This highlights the complexity of HMO influences on infant growth, suggesting that different HMOs may exert distinct metabolic effects depending on their structure and interactions with other milk bioactive components.

Among the sialylated HMOs, our findings align with previous research reporting a positive association between 3’SL and WAZ, consistent with other studies (4,14,38,42,43). Additionally, LSTb was positively associated with WAZ in Saben’s study, reinforcing the potential role of specific sialylated structures in supporting weight gain (43). In contrast, Tonon et al. (2019) found a negative association between 6’SL and WAZ, indicating that not all sialylated HMOs function similarly in growth regulation (23). These divergent findings suggest that the metabolic effects of sialylated HMOs may be structure-dependent.

Notably, the positive growth outcomes observed in our study occurred in a population with an average breastfeeding duration of 69 weeks. This suggests that both early milk composition—reflected in HMO concentrations—and sustained breastfeeding contribute to optimal growth. However, the stability of HMO associations after weighing all children breastfeeding at 6 and 24 months indicates that breastfeeding duration itself may not be a primary driver of these associations.

One limitation of our study is that we did not measure or estimate the total volume of human milk consumed over the first year of life. As a result, we may not fully capture the absolute HMO intake and its effect on growth. Additionally, we measured 19 HMO concentrations at only one time point (one month postpartum). While this presents a limitation, prior evidence suggests that some HMOs, including 2’FL, LSTb, and DSLNT remain stable throughout lactation (33), supporting the reliability of our measurements.

These considerations are particularly relevant given the mixed effects observed in Fig 3, where estimates for 3’SL, LSTb, and DFLac were weakly positive but not statistically significant. In contrast, FLNH exhibited a significant negative association with WAZ across all models (Panels A–C), even after IPCW adjustments, reinforcing a potential inverse relationship between early FLNH levels and later growth outcomes.

These findings highlight the complex interplay between HMO composition, intake levels, and infant growth trajectories, underscoring the need for further research into the mechanisms underlying these associations.

In terms of WLZ, our analysis revealed that multiple HMOs, including DSLNH, FLNH and LNH, were negatively associated with this growth indicator across regression models. These findings may reflect stronger positive associations with LAZ compared to WAZ, leading to a reduced weight-for-length ratio. This aligns with Jorgensen et al. (2020), who also reported an inverse association between FLNH and WLZ (25). It is possible that these associations may reflect FLNH’s role in promoting a more diverse gut microbiota, which could regulate metabolism and prevent excessive weight gain (44).

Analyses weighted breastfeeding at 6 and 24-month time points were performed to explore the impact of breastfeeding duration on the association between HMOs and growth. Minimal changes observed in the outcomes when comparing models with and without weighting suggest that breastfeeding duration does not dramatically change the associations. The role of HMOs on the microbiome is well-documented, as they serve as prebiotics that selectively promote the growth of beneficial gut bacteria. This effect was observed for all children in the cohort, not just those who breastfed for the full 2 years, suggesting that HMO-driven microbiome modulation occurs even among children who were breastfed for a brief period in early life (11,45).

In the sensitivity analysis (S1-4 Fig), we performed residual (centering) adjustment analysis to account for potential multicollinearity, as described in the statistical analysis section. After re-running the three models with these adjustments, the results demonstrated no substantial changes in the estimates or the 95% CIs compared to the main findings. These results reinforce the reliability of our main findings and support the conclusion that each HMO’s effect is independent of intra-group correlations.

Our findings highlight the complex and complementary roles of HMOs in regulating growth patterns, with some promoting balanced growth while others limit disproportionate weight gain. This underscores the importance of considering multiple growth indicators to understand the nuanced effects of HMOs on growth and nutritional status.

Our findings should be interpreted in light of the study’s exploratory design. Because human milk oligosaccharides are structurally interrelated and biologically correlated, applying strict multiple-comparison corrections would have markedly reduced statistical power and increased the risk of Type II errors, potentially obscuring meaningful biological associations. Therefore, we emphasized effect sizes, confidence intervals, and the consistency of observed patterns rather than relying solely on corrected p-values to guide interpretation.

The generalizability of our findings is primarily influenced by the characteristics of the study population and setting. Our study was conducted within a population-based cohort from León, Nicaragua, a context that may differ from other geographic, socioeconomic, and nutritional environments. While the biological mechanisms by which HMOs influence growth patterns are likely to be similar across human populations, variations in maternal health, infant feeding practices, socioeconomic conditions, and environmental exposures may limit the direct applicability of our findings to other settings, particularly high-income countries or regions with different breastfeeding patterns.

While we hypothesize that HMOs influence growth through gut microbiota modulation, this study lacks microbiome data to validate these claims. Future studies should incorporate microbiome analyses and interventional trials with HMO supplementation to establish causation.

## Conclusion

Our findings indicate that specific HMOs measured early in lactation are associated with distinct growth outcomes at 24 months, supporting their structure-specific roles in promoting linear growth. Despite limitations such as single-time-point measurement and unmeasured milk intake, the consistency of associations and alignment with prior evidence strengthen the validity of our results. These insights from a Central American cohort underscore the need for mechanistic studies and intervention trials in LMIC to address early growth deficits.

## Author contributions

The authors’ responsibilities were as follows – SV, NV, FB, SBD, LZ: designed the research; FG, LG, CTR, LB: conducted the research; LZ: analyzed the data; LZ: wrote the manuscript; LZ: had responsibility for the final content; RR, SV, NV, SME, JKE, SBD: provided guidance on study design, data analysis, and interpretation of results.

## Data Availability

The data underlying the results presented in the study contain de-identified human participant information. De-identified data may be made available upon reasonable request to the corresponding author, subject to approval by the relevant Institutional Review Board and data use agreements.

## Acknowledgements

We gratefully acknowledge the SAGE field team for their dedicated efforts in collecting human milk samples, epidemiologic data, and infant length and weight measurements. No new raw data was collected for this study.

## Supporting information

**S1 Fig.** Spearman correlation matrix of human milk oligosaccharide (HMO) concentrations before residual adjustment (RA).

**S2 Fig.** Association between Human Milk Oligosaccharide (HMO) concentrations at one month postpartum and length-for-age Z scores at 24 months of age, after residual centering adjustment. (β coefficients and 95% CIs).

**S3 Fig.** Association between Human Milk Oligosaccharide (HMO) concentrations at one month postpartum and weight-for-age Z scores at 24 months of age, after residual centering adjustment. (β coefficients and 95% CIs).

**S4 Fig.** Association between Human Milk Oligosaccharide (HMO) concentrations at one month postpartum and weight-for-length Z scores at 24 months of age, after residual centering adjustment. (β coefficients and 95% CIs).

## Notes

### Competing Interest Statement

The authors have declared no competing interest.

### Funding Statement

The author(s) received no specific funding for this work.

